# Correcting excess mortality for pandemic-associated population decreases

**DOI:** 10.1101/2021.02.10.21251461

**Authors:** Jeremy Samuel Faust, Chengan Du, Shu-Xia Li, Zhenqiu Lin, Harlan M. Krumholz

## Abstract

**Objectives:** We identify a correction for excess mortality that takes the sudden unexpected changes in the size of the United States population into account.

**Design:** This is a weekly cross-sectional analysis of all-cause mortality since week 5, 2020. We describe and apply a simple correction that takes population changes into account in order to provide corrected weekly estimates of expected deaths for 2020 and 2021.

**Setting:** The United States.

**Participants:** All United States residents.

**Interventions:** The covid-19 pandemic.

**Main outcome measures:** Expected and excess mortality for the United States during the covid-19 period.

**Results:** As of week 53, 2020 (ending January 2, 2021), approximately >10,200 more excess deaths have occurred in the United States than could be detected if expected deaths projections were not amended to reflect population decreases during 2020. The figure is projected to rise to >12,600 (>600 weekly) by week 5, 2021. Assuming recent excess mortality and pandemic-associated visa reductions continue until the earliest time herd immunity could be approached resulting from a combination of infections and vaccinations (week 17, 2021), if point estimates of expected deaths are not corrected, expected deaths will be overestimated (and therefore potential excess mortality underestimated) by ∼43,000 during 2021, or >53,300 since the outbreak of the pandemic measurement period (beginning week 5, 2020). By late December 2021, weekly expected death differences are projected to approach 1,000 per week.

**Conclusions:** Current models measuring excess mortality should be revised immediately so that public health officials do not lose the ability to detect ongoing excess mortality as the population changes continue to compound, lowering the number of weekly expected deaths. A similar approach should be used in the middle and late phases of all future pandemics.

## Introduction

In a pandemic, the ability to detect excess mortality is paramount to policy decision-making. Detection of excess mortality relies on accurate mortality expectations. Unfortunately, covid-19 has corrupted critical assumptions upon which usual predictive models rely. Specifically, there are currently fewer permanent United States residents than projected before the pandemic, owing to excess mortality and decreases in inward migration.

Current Centers for Disease Control and Prevention and other published mortality models do not take real-time population changes into account, assuming that established population trends have not deviated substantially, and therefore minimally affect the accuracy of expected death projections.^1–4^ Sudden changes in the United States population size during 2020 renders that assumption inadequate.

Excess mortality will be underestimated if expected death projections are not lowered to account for these population changes. The cumulative effect of this substantially blunts the ability to detect excess mortality during periods in which mortality rates only modestly exceed projected expectations. Moreover, the insidious compounding of these effects continuously decreases the population denominator, thereby further evading detection.

Here, we describe a simple correction procedure to improve the estimate of excess mortality during the 2020 covid-19 pandemic period and show its effect. We then conservatively project the anticipated magnitude of effect that is likely to occur during 2021 unless the technique we describe is widely adopted. This approach has widespread applicability to future excess death estimates when population sizes are in flux.

## Methods

We performed a cross-sectional analysis of weekly excess deaths in the United States during the covid-19 pandemic period.

First, we determined excess deaths, defined as the difference between observed and expected deaths, using a “standard” approach.^5,6^ For granularity and accuracy, the United States population was divided into 6 age brackets and further divided by sex for a total of 12 demographic groups as follows: 0-24 female, 0-24 male, 25-44 female, 25-44 male, 45-64 female, 45-64 male, 65-74 female, 65-74 male, 75-84 female, 75-84 male, ≥85 female, and ≥85 male.

For observed mortality we used weekly figures provided by the National Center for Health Statistics at the United States Centers for Disease Control and Prevention. For weeks 6-53 2020 (with week 53 ending January 2^nd^, 2021, as per the CDC), published weighted figures were used in order to minimize known lag effects.^7^

To calculate standard expected deaths, we considered ongoing changes due to normal population growth and aging on a weekly basis, as elsewhere.^5^ Specifically, we applied seasonal autoregressive integrated moving average (ARIMA) to project the weekly population for all 12 groups for 2020-2021 using the data from the US Centers for Disease Control and Prevention for 2014-2019.^8,9^ The seasonal period was chosen at 52 weeks to project weekly death expectations for 2020 and 2021 for all 12 groups. We then summed the estimated contributions from the groups to obtain a point estimate of the total number of expected weekly deaths for the United States from week 6 of 2020 through Week 52 of 2021. Both 80% and 95% confidence interval estimates were determined by combining the confidence interval from the 12 groups, assuming independence among groups (see Tables S4-S16).

Next, for each of the 12 groups, we calculated “corrected” weekly estimates for expected deaths taking pandemic-associated changes in population into effect. To do this, weekly all-cause mortality incident rates using point estimates calculated via the standard approach above were first determined. Then, for each week, we calculated a revised estimated population for each of the 12 groups by subtracting the cumulative number of excess deaths during the covid-19 pandemic to that point and the cumulative decrease in permanent resident visas granted to persons arriving from abroad (owing to official policies halting such movements) for that group from the corresponding original population projection.^9^

The decrease in permanent resident visas granted during 2020 was determined by subtracting the month-over-month difference between 2020 and 2019 data, as reported by the United States Department of State (Table S1; see Supplemental Appendix Notes).^10^ Monthly figures were divided by 4 to create weekly estimates. We then apportioned the resulting difference among the 12 demographic groups according to the demographic breakdown of visas granted in 2019 (Table S2).^11^ Finally, to calculate the corrected expected number of weekly deaths, the previously determined all-cause mortality incident rate was multiplied by the revised population estimate (i.e. the corrected denominator).

The mathematical description of this procedure is provided as follows:

We assume there were 53 weeks during 2020. For week 6 < *i* ≤ 53, we defined the original projected population for each of the 12 demographics (denoted by k, where 1 ≤ *k* ≤ 12) in week i of 2020 to be *n*_*i,k*_; the original point estimate for all-cause deaths to be 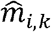; the sum of all previous weeks’ excess death to be *P*_*i*-1,*k*_ (starting from week 6, 2020); the sum of all previous weeks’ decreases in inward immigration to be *Q*_*i*-1,*k*_ (starting from week 6, 2020). Therefore, the corrected expected deaths for week *i* (since week 6, 2020) after correction 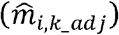 satisfies:

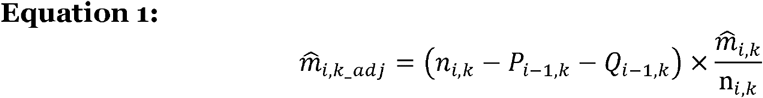

The above equation also holds for the weeks in 2021, where *P*_*i* -1,*k*_ and *Q*_*i* -1,*k*_ denote the sum of all previous weeks’ excess death and the sum of all previous weeks’ decreases in inward immigration since week 6, 2020.

In order to project expected deaths for 2021, we assumed that pandemic-related population changes would continue through week 17 and cease at that point (the week ending May 1, 2021). For simplicity, excess deaths for week 1-17 of 2021 were estimated using the provisional mean excess mortality for each of the 12 demographics reported for weeks 49-52, 2020 (see Tables S4-S16, orange text), though these data remain incomplete and CDC models suggest this assumption will underestimate mean excess mortality during this time.^12^ To estimate ongoing decreases in permanent visas for these weeks, we conservatively assumed a decrease 0f 70% for weeks 1-17 of 2021, although the mean during the pandemic period through December 2020 was actually >80% (Supplemental Appendix Notes, Table S3, and Tables S4-S16, orange text).^9,10^ Expected and excess deaths were reported to the hundreds place in the text and to the ones place in tables and figures.

This study was not subject to institutional review because it relied solely upon data available to the public.

All the analysis was done through R version 4.0.2 and Microsoft Excel version 16.44.

## Results

Our study included 3,079,937 all-cause deaths in the United States from week 5, 2020 through week 53 of 2020, based on preliminary CDC/NCHS data as of February 4, 2021.^7^2/10/2021 1:03:00 PM Absent the covid-19 pandemic, we estimated that the United States population would have increased from approximately 329,525,900 in week 5, 2020 to 331,381,200 by week 53, 2020 residents during the study period (Table 1). Based on standard calculations that take this growth into account, approximately 438,200 excess deaths occurred during that period (lower 95% confidence boundary 282,400) based on preliminary data. However, as of week 53 2020, we estimate that the United States population was actually 330,624,000 persons (757,200 fewer than originally projected), owing to the noted pandemic-associated excess mortality and a measured decrease in the number new permanent visa-granted residents arriving to the United States (approximately 316,300 persons) since February 2020 (Table 1, Table S1). As a result of this denominator change, as of week 53, 2020 we measured that approximately >10,200 more excess deaths occurred in the United States in 2020 than detectable using standard uncorrected approaches (Figure 1).

**Table 1.** Weekly population, calculated expected, observed, excess deaths, and inward migration related population changes, United States, Week 5, 2020 through Week 52, 2021.

| Week ending | Week # | Weekly population | Expected deaths (point estimate) | Lo.80 | Hi.80 | Lo.95 | Hi.95 | Observed deaths (weighted) | Excess deaths | Excess deaths (cumulative) | Decrease in usual inward migration | Decrease in usual inward migration (cumulative) | Corrected population (original-excess deaths-decreased migration) |
| --- | --- | --- | --- | --- | --- | --- | --- | --- | --- | --- | --- | --- | --- |
| 2/8/20 | Week 6 | 329,525,873 | 59,403 | 58,799 | 60,007 | 58,479 | 60,327 | 59,369 | -34 | -34 | -737 | -737 | 329,525,873 |
| 2/15/20 | Week 7 | 329,566,071 | 58,895 | 58,185 | 59,606 | 57,650 | 60,141 | 58,783 | -112 | -146 | -737 | -1,474 | 329,566,842 |
| 2/22/20 | Week 8 | 329,606,270 | 58,199 | 57,375 | 59,023 | 56,568 | 59,830 | 58,866 | 667 | 521 | -737 | -2,211 | 329,607,153 |
| 2/29/20 | Week 9 | 329,646,468 | 57,708 | 56,801 | 58,616 | 55,719 | 59,698 | 59,274 | 1,566 | 2,087 | -737 | -2,948 | 329,646,684 |
| 3/7/20 | Week 10 | 329,686,667 | 58,422 | 57,456 | 59,387 | 56,624 | 60,219 | 59,643 | 1,221 | 3,308 | 3,309 | 361 | 329,685,317 |
| 3/14/20 | Week 11 | 329,726,865 | 57,858 | 56,849 | 58,868 | 55,844 | 59,872 | 58,637 | 779 | 4,087 | 3,309 | 3,670 | 329,720,249 |
| 3/21/20 | Week 12 | 329,767,063 | 57,296 | 56,255 | 58,336 | 55,088 | 59,504 | 59,171 | 1,875 | 5,962 | 3,309 | 6,978 | 329,759,668 |
| 3/28/20 | Week 13 | 329,807,262 | 56,723 | 55,660 | 57,786 | 54,291 | 59,154 | 62,972 | 6,249 | 12,211 | 3,309 | 10,287 | 329,794,683 |
| 4/4/20 | Week 14 | 329,847,460 | 56,865 | 55,786 | 57,943 | 54,354 | 59,376 | 72,235 | 15,370 | 27,581 | 9,263 | 19,550 | 329,825,323 |
| 4/11/20 | Week 15 | 329,887,659 | 56,261 | 55,172 | 57,351 | 53,518 | 59,005 | 79,020 | 22,759 | 50,340 | 9,263 | 28,813 | 329,840,888 |
| 4/18/20 | Week 16 | 329,927,857 | 55,303 | 54,206 | 56,400 | 52,213 | 58,392 | 76,699 | 21,396 | 71,736 | 9,263 | 38,076 | 329,849,065 |
| 4/25/20 | Week 17 | 329,968,056 | 54,721 | 53,619 | 55,823 | 51,354 | 58,088 | 73,831 | 19,110 | 90,846 | 9,263 | 47,339 | 329,858,604 |
| 5/2/20 | Week 18 | 330,008,254 | 54,814 | 53,708 | 55,920 | 51,451 | 58,177 | 69,236 | 14,422 | 105,268 | 9,674 | 57,013 | 329,870,430 |
| 5/9/20 | Week 19 | 330,048,453 | 53,785 | 52,677 | 54,893 | 49,986 | 57,584 | 66,767 | 12,982 | 118,250 | 9,674 | 66,687 | 329,886,532 |
| 5/16/20 | Week 20 | 330,088,651 | 53,398 | 52,288 | 54,508 | 49,512 | 57,284 | 64,429 | 11,031 | 129,281 | 9,674 | 76,361 | 329,904,075 |
| 5/23/20 | Week 21 | 330,128,849 | 53,533 | 52,422 | 54,644 | 49,691 | 57,375 | 61,564 | 8,031 | 137,312 | 9,674 | 86,035 | 329,923,568 |
| 5/30/20 | Week 22 | 330,169,048 | 52,934 | 51,822 | 54,046 | 48,803 | 57,065 | 59,613 | 6,679 | 143,991 | 9,674 | 95,709 | 329,946,062 |
| 6/6/20 | Week 23 | 330,209,246 | 53,592 | 52,479 | 54,704 | 49,691 | 57,492 | 58,828 | 5,236 | 149,227 | 9,241 | 104,950 | 329,969,907 |
| 6/13/20 | Week 24 | 330,249,445 | 53,116 | 52,002 | 54,229 | 49,070 | 57,162 | 57,963 | 4,847 | 154,074 | 9,241 | 114,190 | 329,995,629 |
| 6/20/20 | Week 25 | 330,289,643 | 52,791 | 51,677 | 53,905 | 48,636 | 56,945 | 57,921 | 5,130 | 159,204 | 9,241 | 123,431 | 330,021,740 |
| 6/27/20 | Week 26 | 330,329,842 | 52,638 | 51,524 | 53,752 | 48,402 | 56,875 | 58,423 | 5,785 | 164,989 | 9,241 | 132,671 | 330,047,567 |
| 7/4/20 | Week 27 | 330,370,040 | 52,997 | 51,883 | 54,112 | 48,803 | 57,192 | 59,744 | 6,747 | 171,736 | 8,789 | 141,460 | 330,072,741 |
| 7/11/20 | Week 28 | 330,408,930 | 52,693 | 51,578 | 53,807 | 48,441 | 56,944 | 61,835 | 9,142 | 180,878 | 8,789 | 150,249 | 330,096,095 |
| 7/18/20 | Week 29 | 330,447,820 | 52,286 | 51,171 | 53,401 | 47,902 | 56,670 | 63,070 | 10,784 | 191,662 | 8,789 | 159,038 | 330,117,054 |
| 7/25/20 | Week 30 | 330,486,710 | 52,067 | 50,952 | 53,183 | 47,593 | 56,542 | 64,127 | 12,060 | 203,722 | 8,789 | 167,827 | 330,136,371 |
| 8/1/20 | Week 31 | 330,525,600 | 52,223 | 51,107 | 53,338 | 47,782 | 56,663 | 64,098 | 11,875 | 215,597 | 8,789 | 176,616 | 330,154,412 |
| 8/8/20 | Week 32 | 330,564,490 | 52,430 | 51,315 | 53,546 | 48,077 | 56,783 | 63,730 | 11,300 | 226,897 | 7,998 | 184,614 | 330,172,638 |
| 8/15/20 | Week 33 | 330,603,380 | 51,855 | 50,739 | 52,971 | 47,313 | 56,398 | 63,820 | 11,965 | 238,862 | 7,998 | 192,611 | 330,192,231 |
| 8/22/20 | Week 34 | 330,642,270 | 51,965 | 50,849 | 53,081 | 47,417 | 56,512 | 62,683 | 10,718 | 249,580 | 7,998 | 200,609 | 330,211,159 |
| 8/29/20 | Week 35 | 330,681,160 | 52,069 | 50,953 | 53,185 | 47,591 | 56,547 | 61,214 | 9,145 | 258,725 | 7,998 | 208,606 | 330,231,333 |
| 9/5/20 | Week 36 | 330,720,051 | 52,197 | 51,081 | 53,313 | 47,796 | 56,598 | 60,274 | 8,077 | 266,802 | 6,145 | 214,751 | 330,253,080 |
| 9/12/20 | Week 37 | 330,758,941 | 52,580 | 51,463 | 53,696 | 48,328 | 56,831 | 59,662 | 7,082 | 273,885 | 6,145 | 220,896 | 330,277,748 |
| 9/19/20 | Week 38 | 330,797,831 | 52,856 | 51,739 | 53,973 | 48,741 | 56,971 | 59,649 | 6,793 | 280,678 | 6,145 | 227,040 | 330,303,411 |
| 9/26/20 | Week 39 | 330,836,721 | 53,226 | 52,109 | 54,343 | 49,215 | 57,237 | 60,376 | 7,150 | 287,828 | 6,145 | 233,185 | 330,329,363 |
| 10/3/20 | Week 40 | 330,875,611 | 53,686 | 52,569 | 54,804 | 49,884 | 57,489 | 59,083 | 5,397 | 293,225 | 7,820 | 241,005 | 330,354,959 |
| 10/10/20 | Week 41 | 330,914,501 | 53,808 | 52,690 | 54,925 | 50,049 | 57,567 | 60,344 | 6,536 | 299,761 | 7,820 | 248,825 | 330,380,632 |
| 10/17/20 | Week 42 | 330,953,391 | 54,925 | 53,808 | 56,043 | 51,598 | 58,253 | 58,865 | 3,940 | 303,700 | 7,820 | 256,644 | 330,405,166 |
| 10/24/20 | Week 43 | 330,992,281 | 55,156 | 54,038 | 56,274 | 51,886 | 58,427 | 60,450 | 5,294 | 308,994 | 7,820 | 264,464 | 330,432,297 |
| 10/31/20 | Week 44 | 331,031,171 | 54,989 | 53,871 | 56,107 | 51,721 | 58,258 | 61,637 | 6,648 | 315,642 | 7,820 | 272,284 | 330,458,074 |
| 11/7/20 | Week 45 | 331,070,061 | 55,968 | 54,849 | 57,086 | 52,994 | 58,942 | 65,143 | 9,175 | 324,817 | 6,327 | 278,611 | 330,482,496 |
| 11/14/20 | Week 46 | 331,108,951 | 56,593 | 55,475 | 57,712 | 53,864 | 59,322 | 66,589 | 9,996 | 334,813 | 6,327 | 284,938 | 330,505,884 |
| 11/21/20 | Week 47 | 331,147,841 | 56,823 | 55,704 | 57,941 | 54,196 | 59,449 | 69,005 | 12,182 | 346,995 | 6,327 | 291,265 | 330,528,451 |
| 11/28/20 | Week 48 | 331,186,731 | 56,639 | 55,521 | 57,758 | 53,975 | 59,303 | 70,466 | 13,827 | 360,822 | 6,327 | 297,592 | 330,548,832 |
| 12/5/20 | Week 49 | 331,225,621 | 57,905 | 56,786 | 59,024 | 55,661 | 60,148 | 74,310 | 16,405 | 377,227 | 7,222 | 304,814 | 330,567,569 |
| 12/12/20 | Week 50 | 331,264,511 | 58,347 | 57,227 | 59,466 | 56,375 | 60,318 | 77,924 | 19,577 | 396,804 | 7,358 | 312,172 | 330,582,831 |
| 12/19/20 | Week 51 | 331,303,401 | 58,310 | 57,191 | 59,430 | 56,263 | 60,358 | 76,769 | 18,459 | 415,263 | 7,358 | 319,530 | 330,594,786 |
| 12/26/20 | Week 52 | 331,342,291 | 58,593 | 57,473 | 59,712 | 56,622 | 60,563 | 74,019 | 15,426 | 430,689 | 7,358 | 326,888 | 330,607,859 |
| 1/2/21 | Week 53 | 331,381,182 | 60,280 | 59,160 | 61,400 | 58,865 | 61,696 | 67,807 | 7,527 | 438,216 | 7,358 | 334,247 | 330,623,965 |

**Figure 1.**
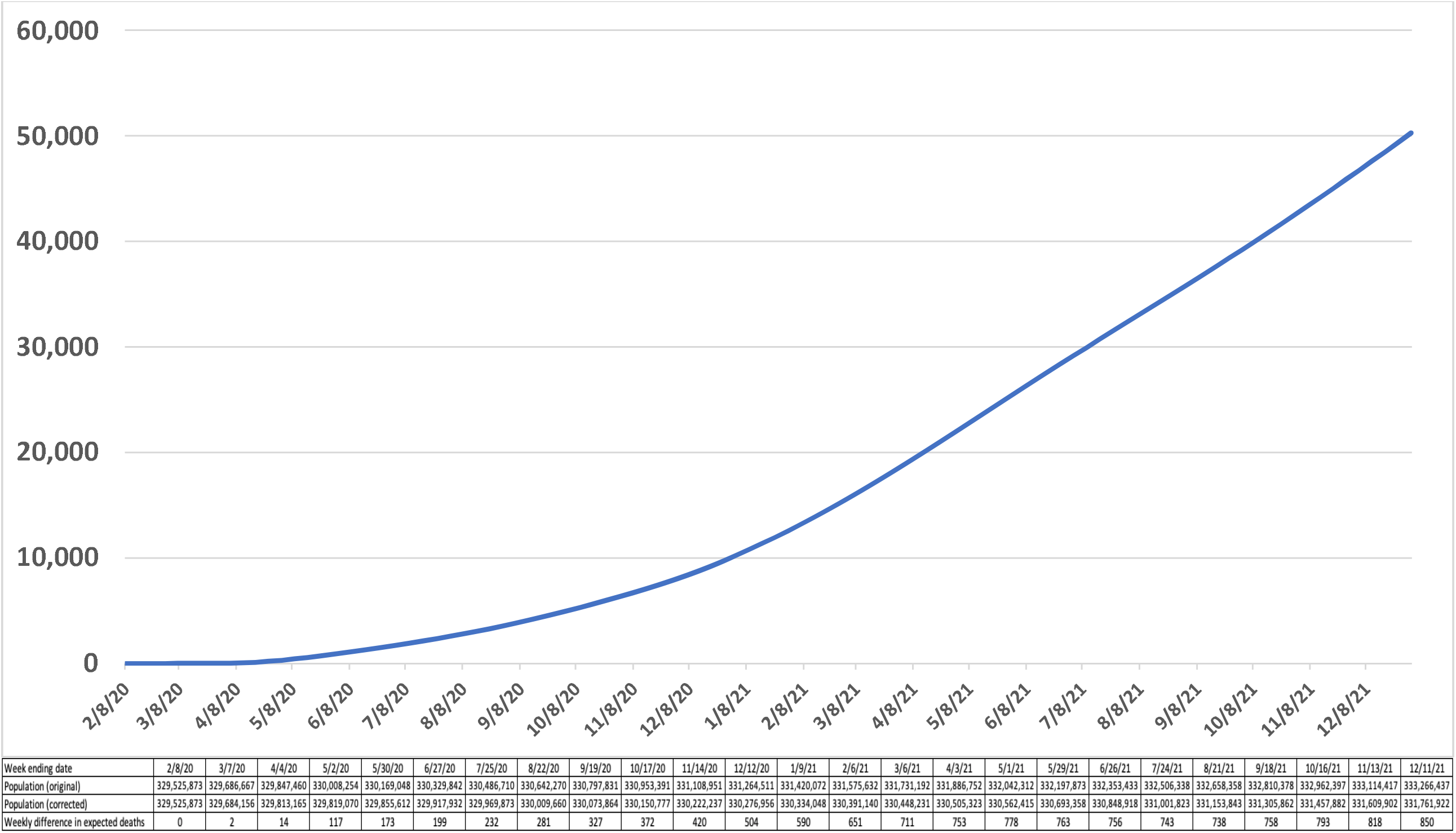
Cumulative difference between corrected and uncorrected expected deaths, United States, week 5, 2020 through week 52, 2021.

Due to the compounding effect, the number of missed excess deaths will rise to >12,600 (>600 weekly) by week 5, 2021 (Figure 1-2). Assuming recent excess mortality and pandemic-associated visa reductions continue until the earliest feasible time herd immunity could be approached resulting from a combination of infections and vaccinations (week 17, 2021), if point estimates of expected deaths are not corrected to reflect these changes, we project expected deaths will be overestimated (and therefore potential excess mortality underestimated) by ∼43,000 during 2021 (Figure 2), or >53,300 since the outbreak of the pandemic measurement period (beginning week 5, 2021). By late December 2021, weekly expected death differences are projected to approach 1,000 per week (∼61,200 expected deaths using the standard calculation versus ∼60,200 using the population-corrected calculation (Figure 3).

**Figure 2.**
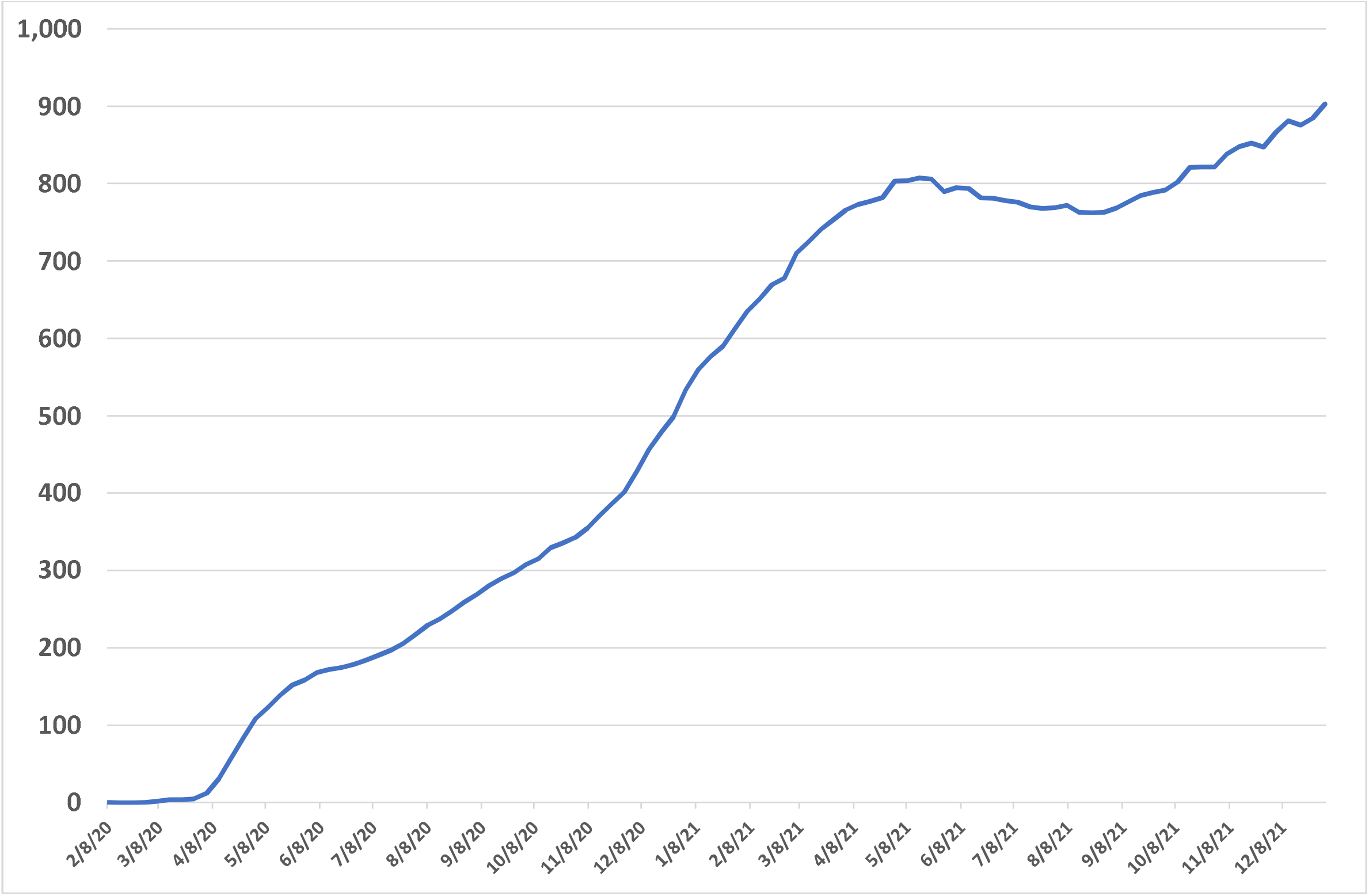
Weekly difference between corrected and uncorrected expected deaths, United States, week 5, 2020 through week 52, 2021.

**Figure 3.**
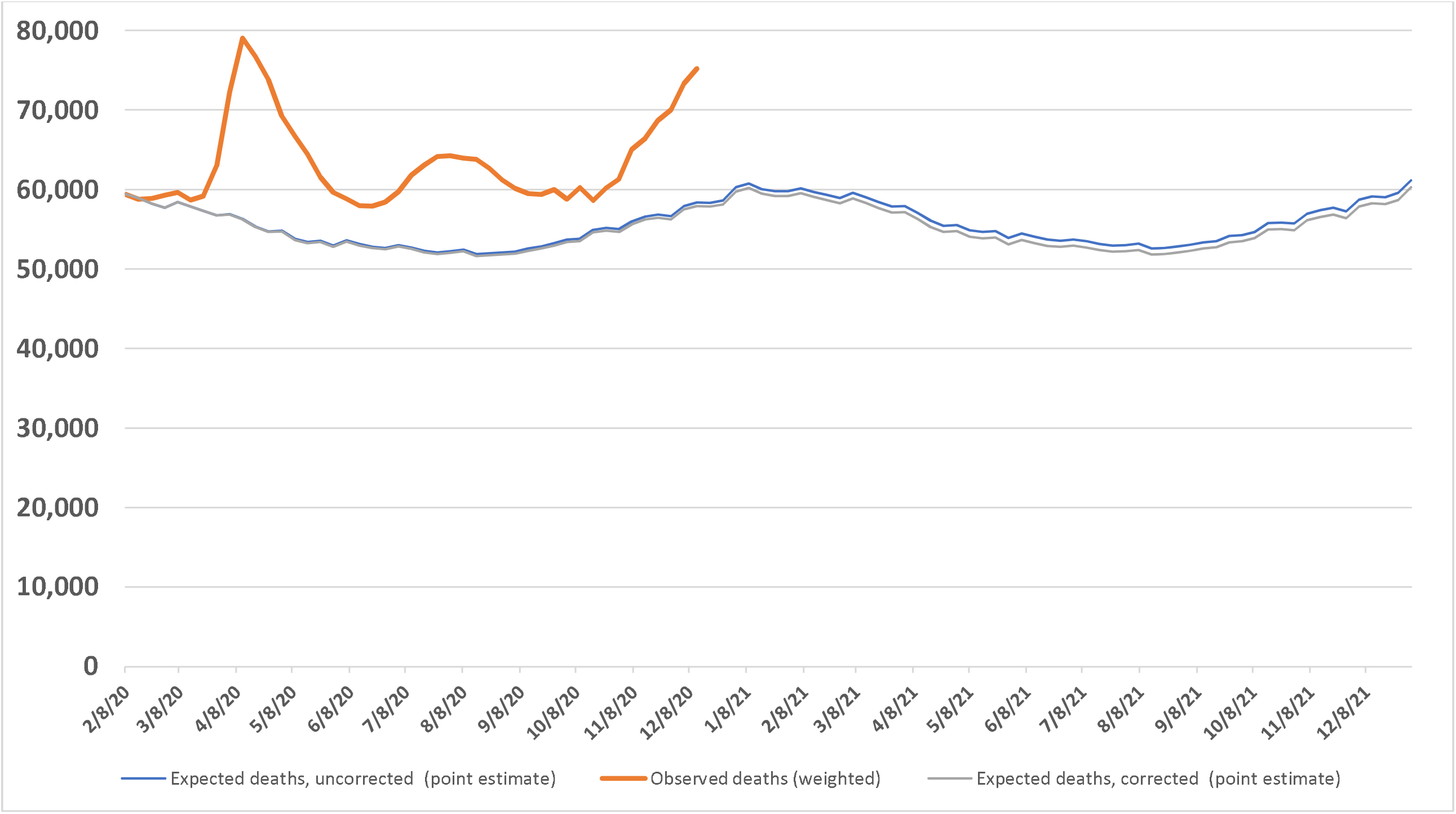
Observed versus uncorrected expected and corrected expected deaths, United States, week 5, 2020 through week 52, 2021.

## Discussion

We find that excess mortality in the United States has been underestimated during the covid-19 pandemic because current models in use do not consider changes in population that have occurred during the crisis. Here, we show that by taking into account such population changes weekly, the number of expected deaths decreases, meaning that excess mortality is greater than would otherwise be detectable.

We also show that this effect is compounding over time such that these quantitative differences will soon translate into an important qualitative reversal. If the measured number of deaths in a coming month is similar to the originally expected deaths estimate, observers will incorrectly conclude that excess mortality is no longer occurring. However, in comparison to the corrected estimate, excess deaths would, in fact, be occurring (Figure 3, the growing area between blue and gray lines). Such a failing to detect ongoing excess death could lead to premature policy changes such as the early curtailing of non-pharmacologic interventions. Moreover, after the pandemic subsides, failure to make the corrections identified here would eventually and misleadingly seem to appear to epidemiologists and public health officials as *deficit mortality* (i.e., measuring fewer deaths than expected). This approach would inaccurately depict a scenario in which unadjusted post-pandemic death rates appear to be lower than at any time in history, when they are not. In addition, because much of the excess mortality that has occurred during the covid-19 pandemic has been in persons with medical comorbidities, the surviving post-pandemic population is likely to collectively have a lower crude all-cause mortality rate than the pre-pandemic population because a smaller proportion of them will have significant comorbidities. Therefore, even the corrected estimates of expected mortality in the future we provide may *still* be overestimated. In fact, our findings obliquely points to the need for a systemic post-pandemic population risk recalibration so that public health professionals can continue to track disease-specific mortality for virtually every cause of death accurately.

A strength of our correction procedure for calculating excess mortality is that it relies upon real data for 2020 (week 5, 2020 to week 53, 2020). A weakness is that we must currently rely upon incomplete provisional data. Therefore, reporting lags render the figures presented to be lower boundary estimates of the effect we have unveiled for 2020, which also artificially lowers our 2021 projections.

An acknowledged weakness in our 2021 projections is that it was necessary to estimate the magnitude of excess mortality and the decrease in permanent visas for weeks 1-17 of 2021. In addition, the assumption that all such pandemic-associated changes would continue until May 1, 2021, and then cease suddenly was arbitrary. However, we believe our approach for this period was conservative; we used average counts from weeks 49-52 of 2020, the most recent for which adequate data is currently available, although the subsequent weeks (January 2021) corresponded to the deadliest weeks of the pandemic in the United States yet recorded.^13^ Thus, our projections for 2021 are almost certain to underestimate the true magnitude of the effect described here. In addition, the changes due to immigration described herein also represent lower boundary estimates, as we considered only permanent visas granted from persons arriving directly from abroad (See Supplemental Appendix Notes). Based on prior years and government travel data (Table S3), approximately twice as many fewer permanent residents likely arrived to the United States during the pandemic period than would be represent by CDC mortality data.

To our knowledge, this report is the first to describe the underestimation of excess mortality resulting from real-time changes in the population during a pandemic or outbreak of any kind. During the covid-19 pandemic, most of the effect we describe resulted from excess mortality rather than immigration. However, in a future pandemic with higher mortality rates among young and middle-aged adults (who comprise a majority of inward migrants, see Table S2), that balance could change. This means that more inclusive and more definitive real-time reporting on the decreases in inward migration might be more important in accurately measuring this effect in future pandemics. In nations with more outward migration than inward migration, the effects described here could have the opposite effect. However, these potential eventualities are built into our procedure (See Equation 1).

We propose that in the middle and late phases of the covid-19 pandemic—and future pandemics that similarly cause substantial and measurable changes in the population size—the correction methodology described here will help officials more accurately detect excess mortality later in the pandemic. Failing to adopt the approach described here is almost certain to lead to the premature declaration that excess mortality has ceased to occur both now and in any similar instances in the future. That the problem we identify here has not been considered in the past is a result of the fact that there has never been an event with this magnitude of excess mortality during an era in which real-time state, national, and even international data became available to epidemiologists and public officials with such a short time lag and on an ongoing basis.

Excess mortality remains one of the most reliable metrics for measuring a pandemic’s effect, as it is agnostic to cause of death. However, to leverage excess death data properly, the ability to detect it later in pandemics must be maintained. Future research will include updated data and assess whether the effects described herein can be reliably applied at the city or state level, where denominators are smaller. In addition, the correction methodology we have identified may be improved upon by further by considering excess mortality in certain risk pools such as among persons living in long term care facilities, or in certain high-risk areas. To ensure our policy decisions are based on the best available evidence, officials should update United States and global expected death estimates immediately.

## Data Availability

The data are available to the public and were available to all authors at all times.

## Acknowledgements

We wish to thank Lauren M. Rossen, PhD, MS (National Center for Health Statistics), for facilitating the public release of National Center for Health Statistics data used for this study, and Julia Gelatt, PhD (Migration Policy Institute) for expertise and help identifying migration data. Neither individual received compensation for their contributions.

## Patient and public involvement statement

It was not appropriate or possible to involve patients or the public in the design, or conduct, or reporting, or dissemination plans of our research.

## Contributor statement

Dr. Faust conceived of the project, designed it, and wrote the manuscript in consultation with Dr. Krumholz. Dr. Du, Li, and Lin provided the statistical modeling framework and planning. Dr. Du performed the primary statistical modeling. analysis. All authors were involved in the analysis of the findings and contributed to the manuscript creation process either via direct writing or editing.

## Supplemental Appendix

See attached pdf.

