## Supplemental methods and tables for "Correcting excess mortality for pandemic-associated population decreases"

This appendix has been provided by the authors to give readers additional information about their work.

**Notes on permanent inward migration to the United States:**

In 2019, 1.03 million persons obtained permanent residency (“green cards”) in the United States.^1^ While some of these visas were granted to persons already on United States soil, it is assumed that the number of persons who arrive on US soil annually is equal to or greater than the number of persons granted green cards in order to maintain this “steady-state.” At the outset of the pandemic period in the United States most inward migration and all permanent residency applications were temporarily halted, and then continued at levels far lower than in previous years (Table S1, Table S3). Nevertheless, in order to be conservative, we elected to consider only changes in the number of persons who obtained lawful permanent resident status visas in the United States arriving directly from foreign soil (Table S1). We then apportioned the difference in visas granted in 2019 and 2020 during the study period according to the demographic breakdown reported in 2019 (Table S2). We feel this approach is conservative as it only considers inward migration among persons already granted permanent visas prior to arrival, changes which were far smaller than the magnitude than the reported decrease in all international travel into the United States during the pandemic period (Table S3). A less conservative estimate would have considered the decrease in persons captured in Table S3 which using figures from 2019 would have been based on 1.03 million arriving permanent residents. Using those figures would have increased our estimate for the number of excess deaths by approximately 10%.

**Table S1:** Permanent resident visas in 2019, 2020, change and 2020 as a share of the 2019 month-over-month figure.

| 2020 as a share of the 2019 monthly average |  |  |  |  |
| --- | --- | --- | --- | --- |
|  | **2019** | **2020** | **Change** | **Percent change** |
| **January** | 39,557 | 43,136 | -3,579 | 112% |
| **February** | 34,710 | 37,658 | -2,948 | 98% |
| **March** | 37,618 | 24,383 | 13,235 | 63% |
| **April** | 38,573 | 1,521 | 37,052 | 4% |
| **May** | 39,393 | 697 | 38,696 | 2% |
| **June** | 38,529 | 1,567 | 36,962 | 4% |
| **July** | 39,568 | 4,412 | 35,156 | 11% |
| **August** | 38,090 | 6,100 | 31,990 | 16% |
| **September** | 39,473 | 14,894 | 24,579 | 39% |
| **October** | 39,966 | 8,687 | 31,279 | 23% |
| **November** | 35,446 | 10,138 | 25,308 | 26% |
| **December** | 40,666 | 11,233 | 29,433 | 29% |
| **March-Dec Average** | 38,517 | 8,044 | 30,473 | 21% |

*Source: U.S. Department of State, Monthly Immigrant Visa Issuance Statistics.*^2^

**Table S2.** Persons Obtaining Lawful Permanent Resident Status by Sex and Age: Fiscal Year 2019.

|  | Female | Male |
| --- | --- | --- |
| <25 | 15.4% | 16.1% |
| 25-44 | 13.2% | 13.5% |
| 45-64 | 13.0% | 12.4% |
| 65-74 | 5.1% | 4.5% |
| 75-84 | 2.7% | 2.1% |
| 85+ | 1.3% | 0.7% |

*Source: collated from Department of Homeland Security, 2019 yearbook.*^3^

**Table S3:** International Passenger Enplanements on U.S. Airlines, Seasonally-Adjusted.

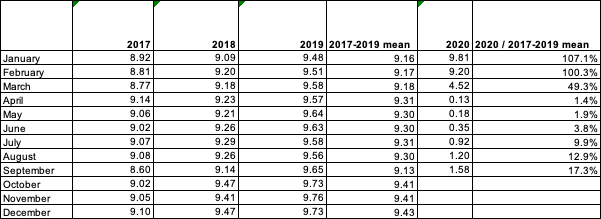

*Source: Bureau of Transportation Statistics, T-100 International Market.*^4^

*Notes: International passenger enplanements (seasonally-adjusted) in millions. Scheduled service only.*

**Table S4.** Weekly population, expected all-cause mortality, excess deaths, population decreases, corrected expected and excess deaths, in US female <25 years, Week 6, 2020 through Week 52, 2021.

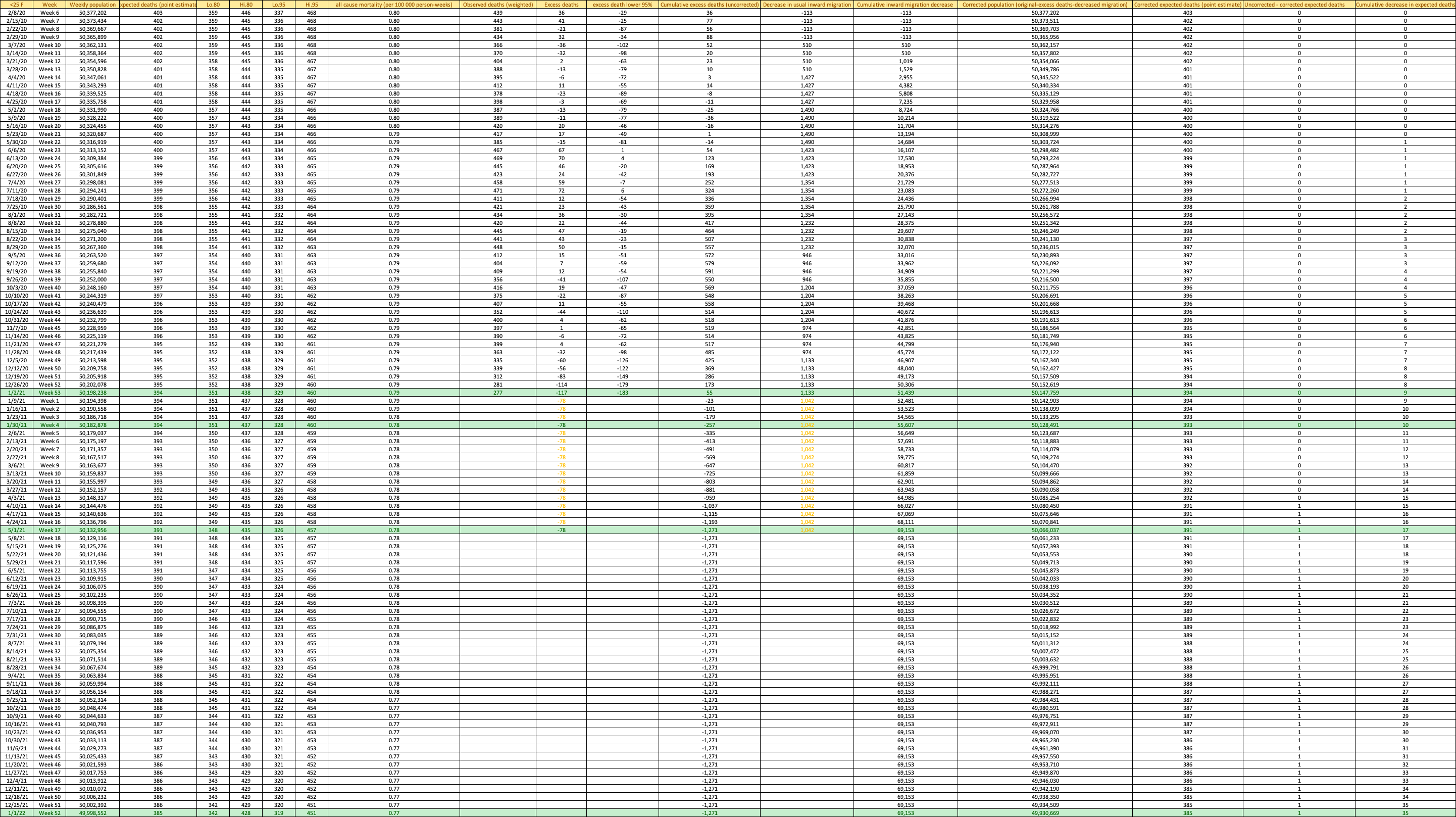

**Table S5.** Weekly population, expected all-cause mortality, excess deaths, population decreases, corrected expected and excess deaths, in US males <25 years, Week 6, 2020 through Week 52, 2021.

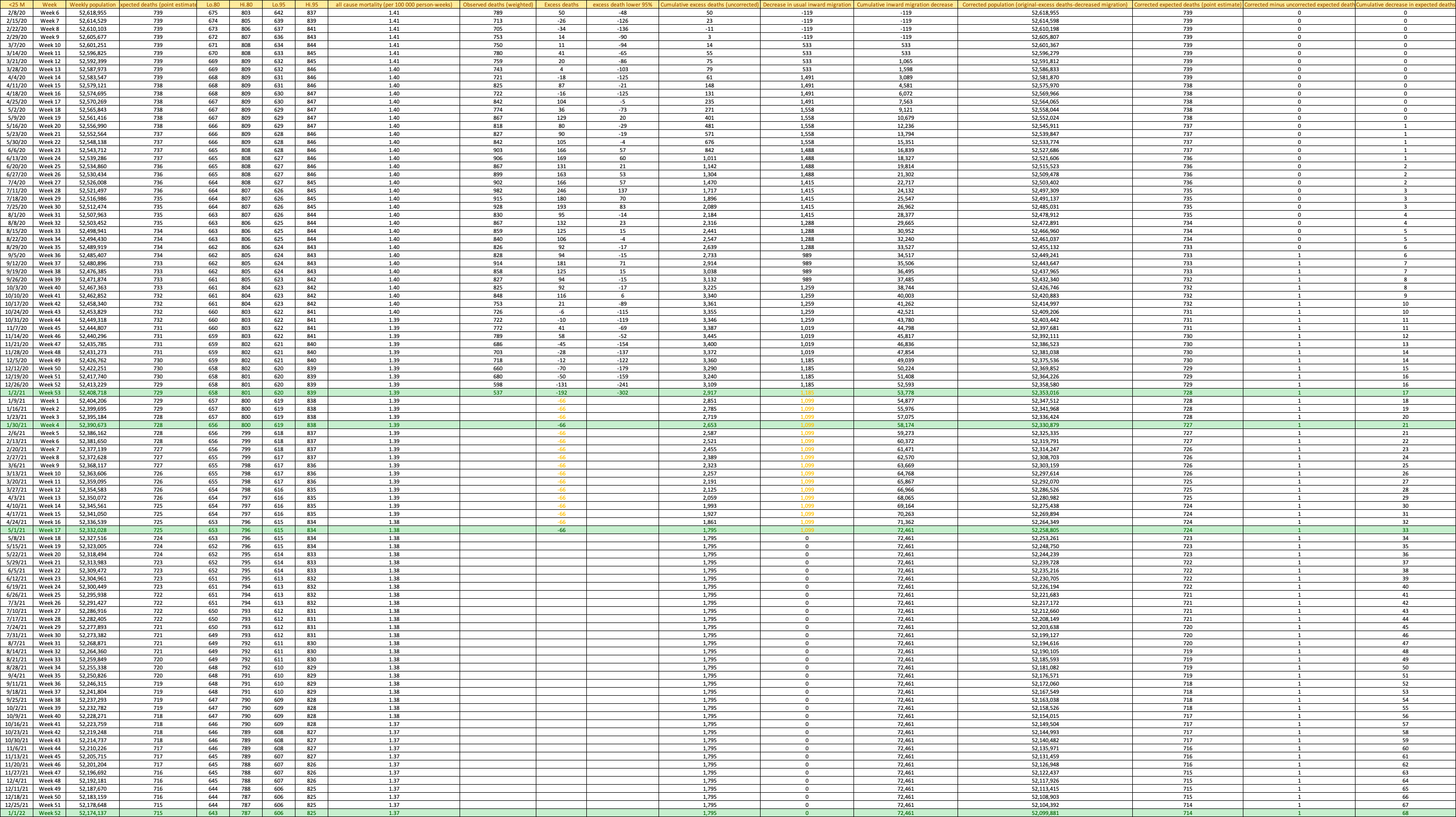

**Table S6.** Weekly population, expected all-cause mortality, excess deaths, population decreases, corrected expected and excess deaths, in US females 25-44 years, Week 6, 2020 through Week 52, 2021.

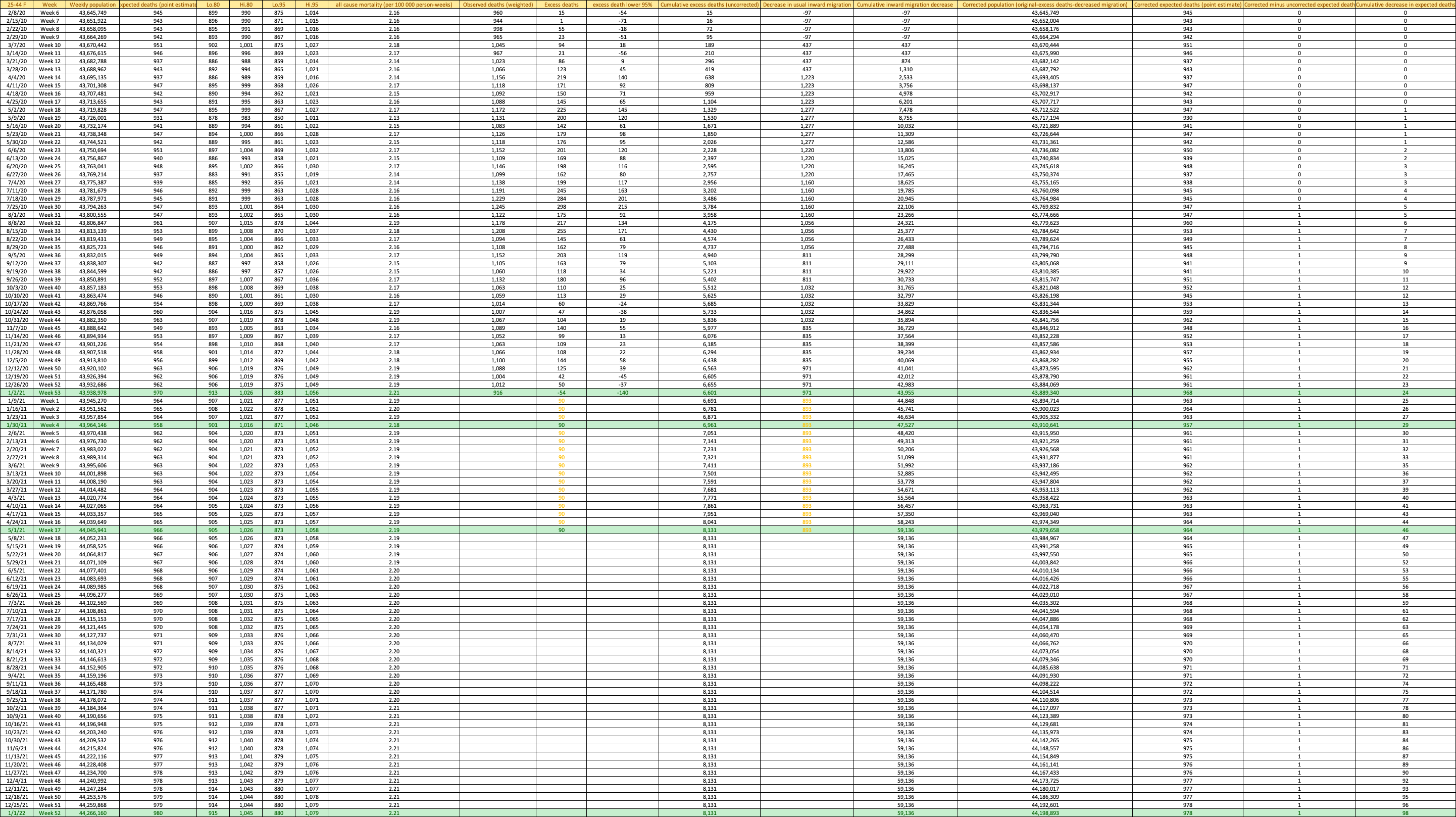

**Table S7.** Weekly population, expected all-cause mortality, excess deaths, population decreases, corrected expected and excess deaths, in US males 25-44 years, Week 6, 2020 through Week 52, 2021.
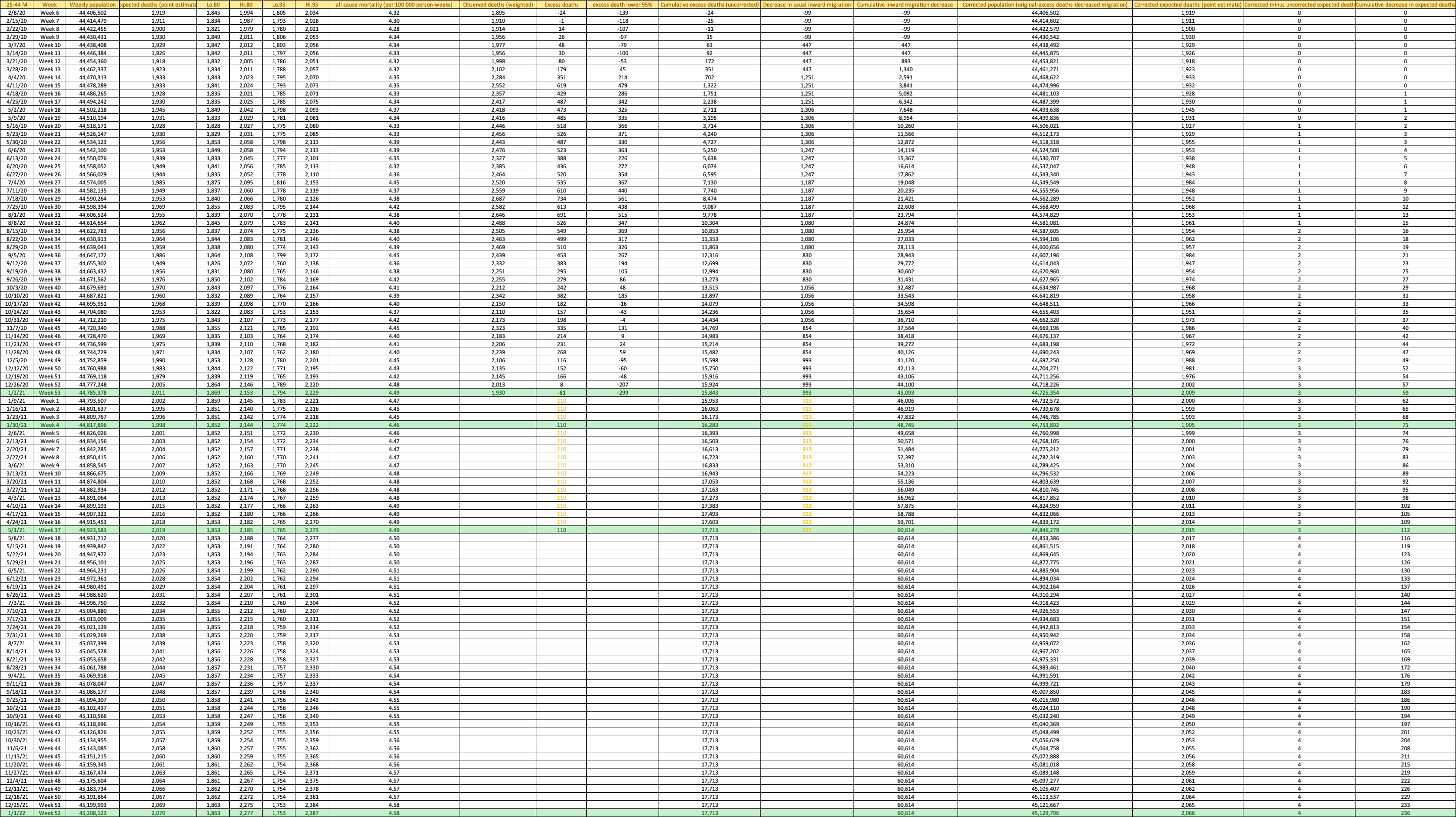

**Table S8.** Weekly population, expected all-cause mortality, excess deaths, population decreases, corrected expected and excess deaths, in US females 45-64 years, Week 6, 2020 through Week 52, 2021.
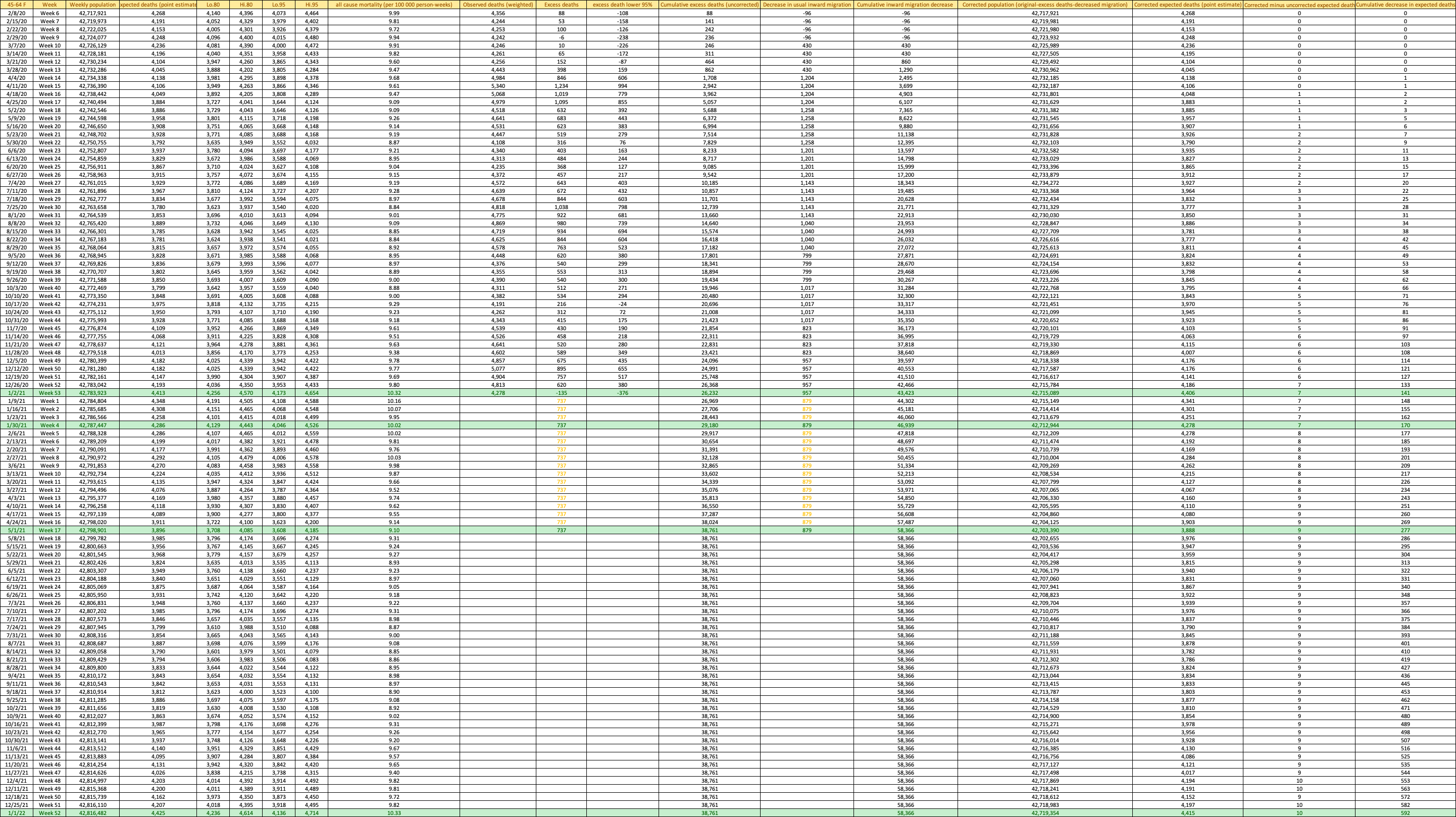

**Table S9.** Weekly population, expected all-cause mortality, excess deaths, population decreases, corrected expected and excess deaths, in US males 45-64 years, Week 6, 2020 through Week 52, 2021.
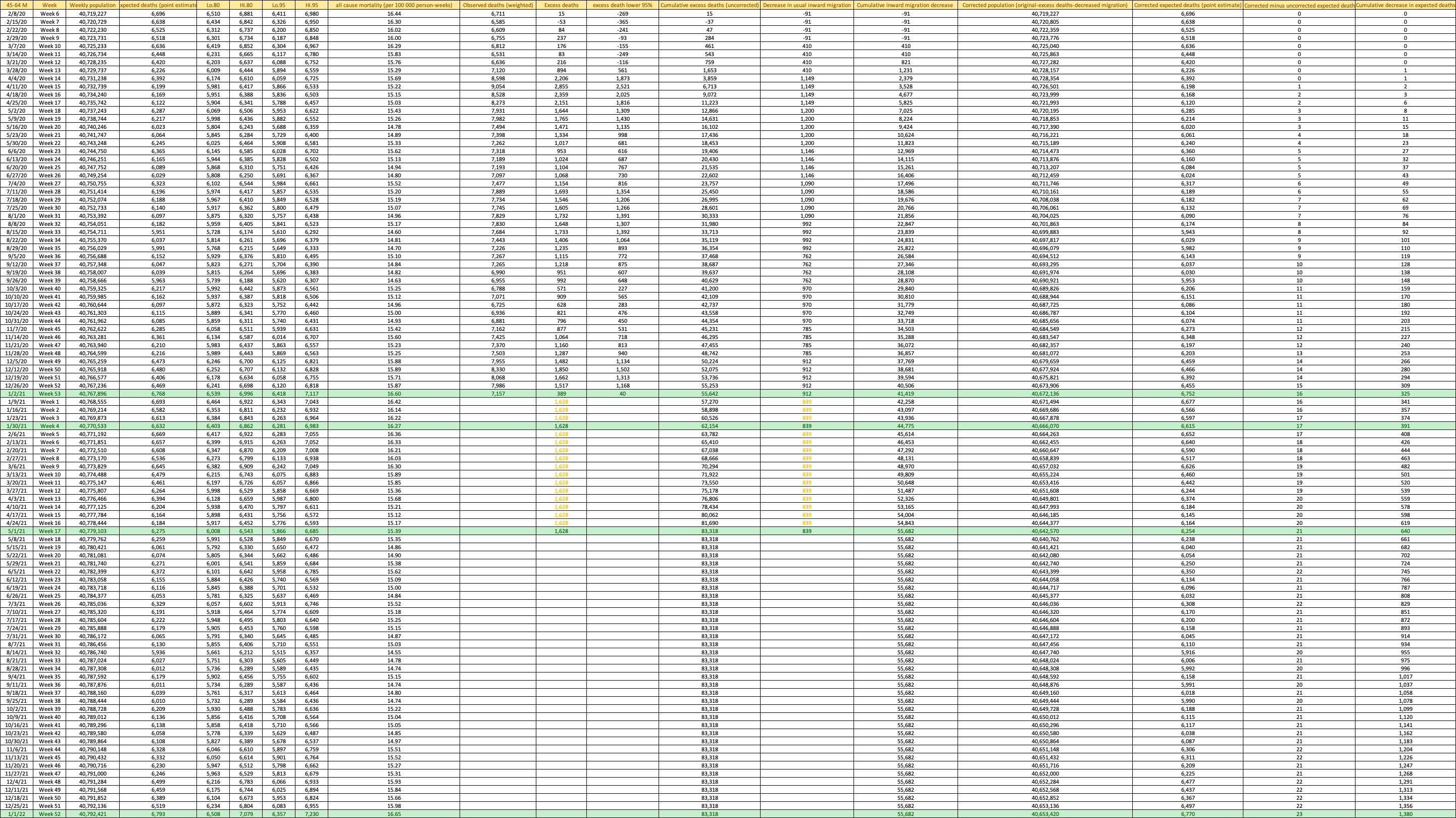

**Table S10.** Weekly population, expected all-cause mortality, excess deaths, population decreases, corrected expected and excess deaths, in US females 65-74 years, Week 6, 2020 through Week 52, 2021.
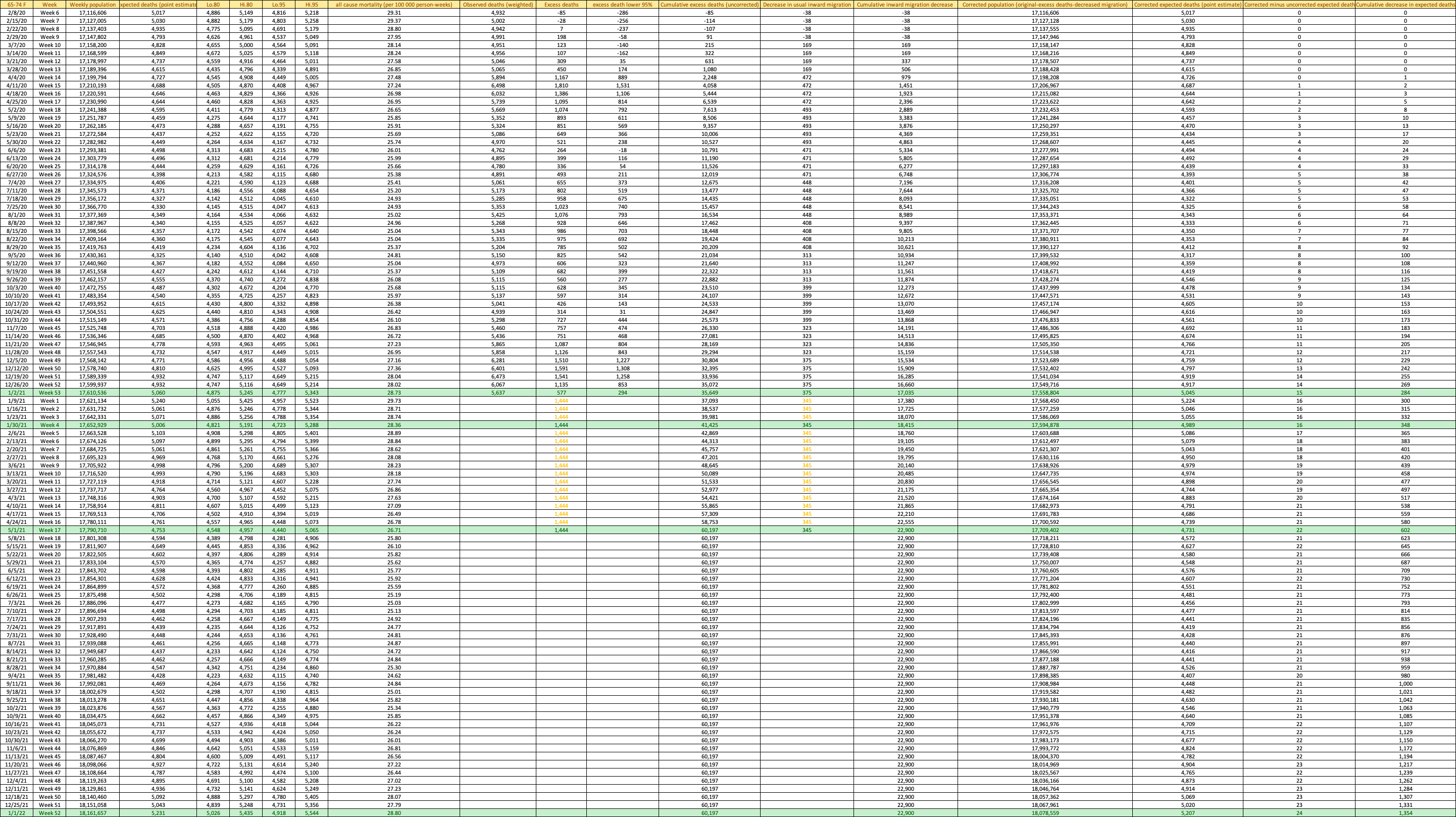

**Table S11.** Weekly population, expected all-cause mortality, excess deaths, population decreases, corrected expected and excess deaths, in US males 65-74 years, Week 6, 2020 through Week 52, 2021.
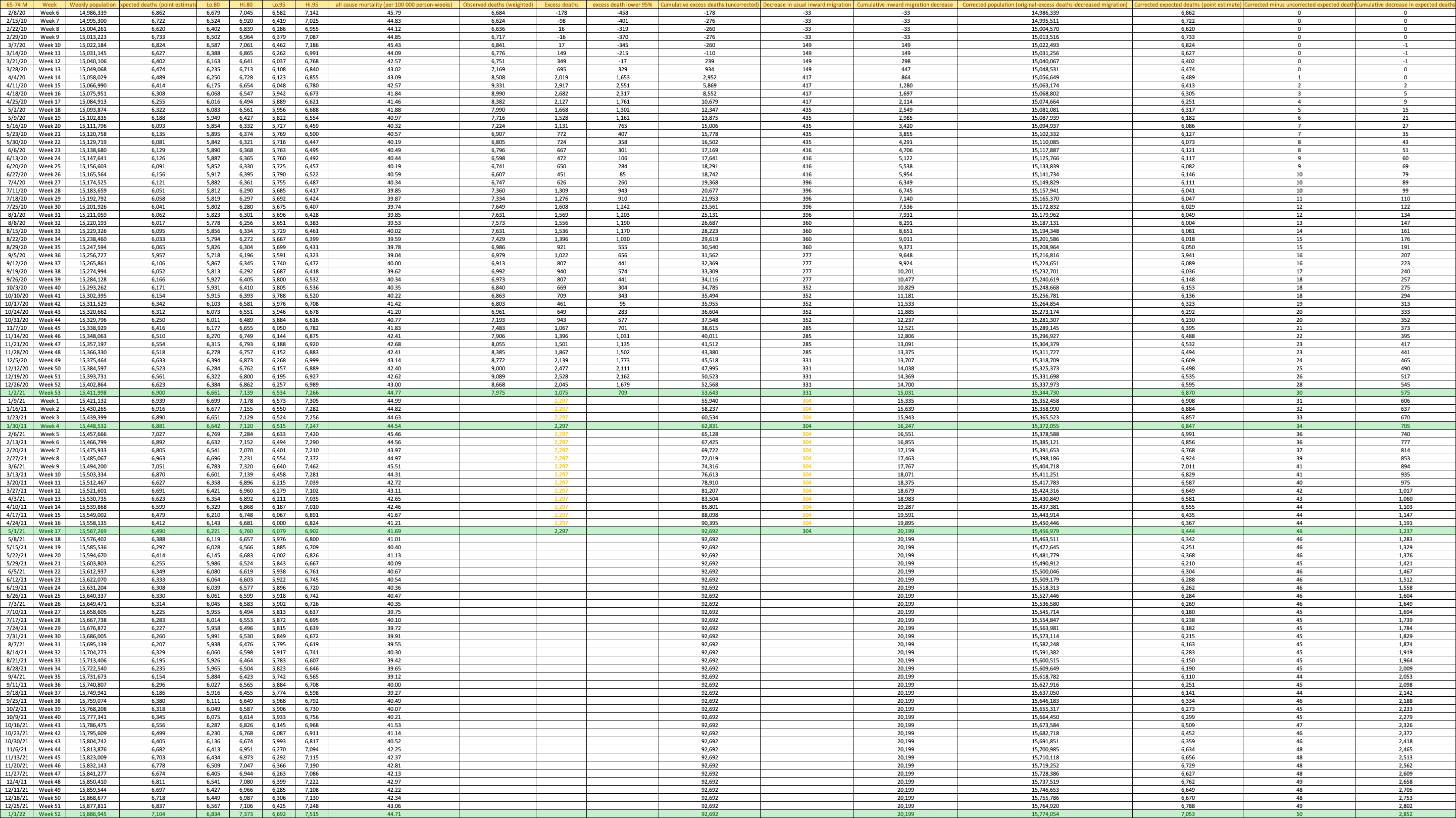

**Table S12.** Weekly population, expected all-cause mortality, excess deaths, population decreases, corrected expected and excess deaths, in US females 75-84 years, Week 6, 2020 through Week 52, 2021.
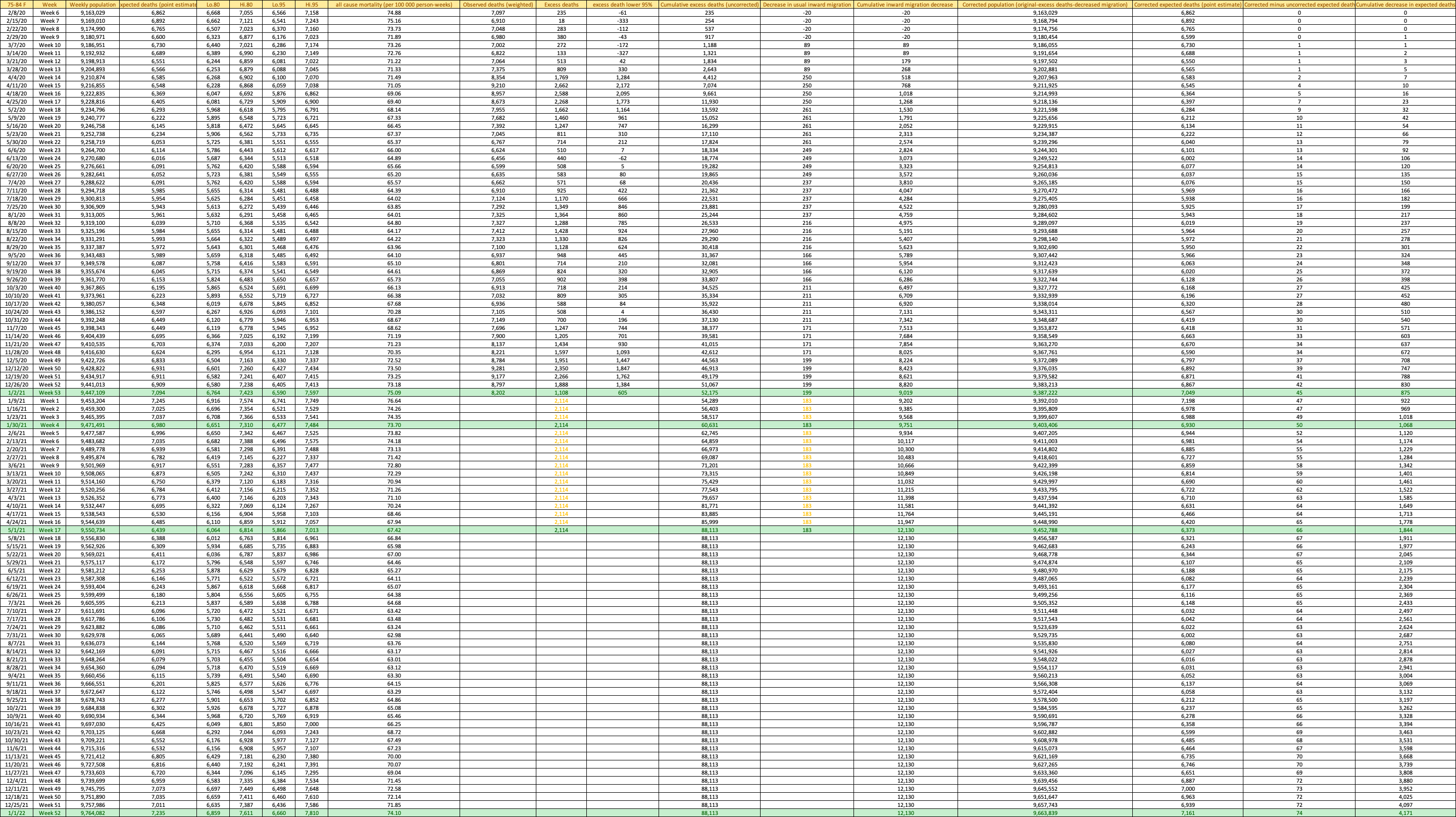
**Table S13.** Weekly population, expected all-cause mortality, excess deaths, population decreases, corrected expected and excess deaths, in US males 75-84 years, Week 6, 2020 through Week 52, 2021.
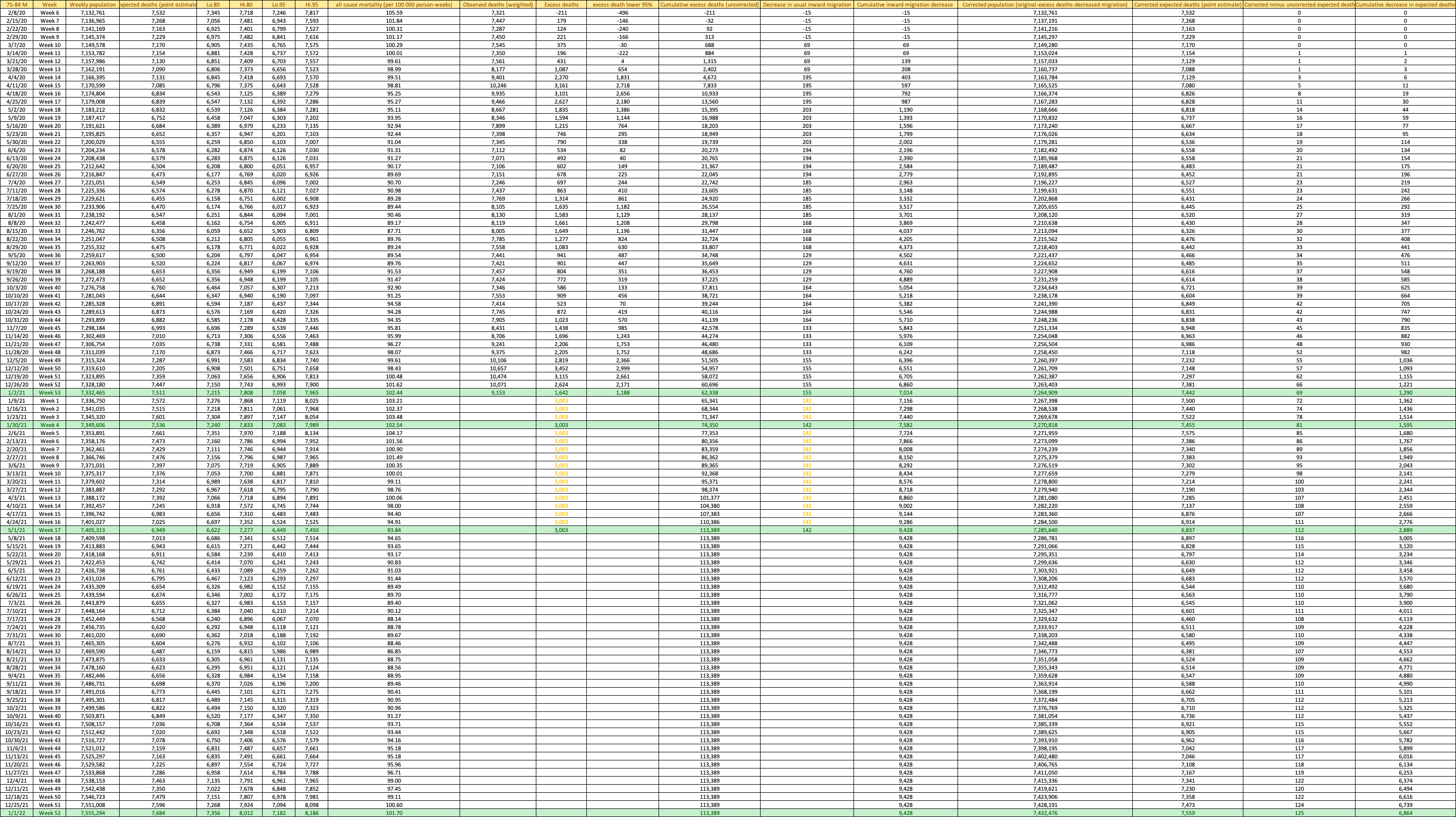

**Table S14.** Weekly population, expected all-cause mortality, excess deaths, population decreases, corrected expected and excess deaths, in US females ≥85 years, Week 6, 2020 through Week 52, 2021.
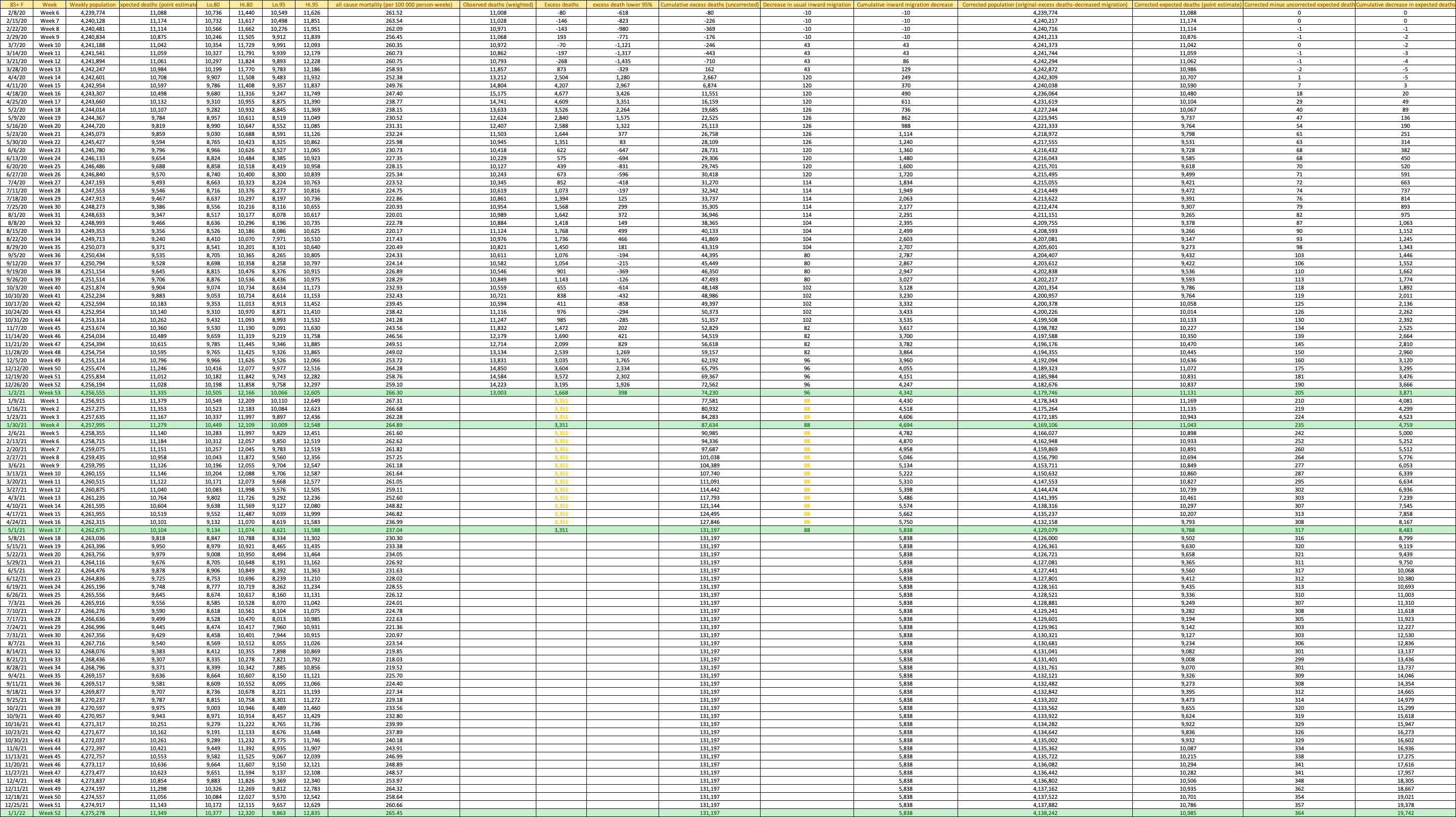
**Table S15.** Weekly population, expected all-cause mortality, excess deaths, population decreases, corrected expected and excess deaths, in US males ≥85 years, Week 6, 2020 through Week 52, 2021.
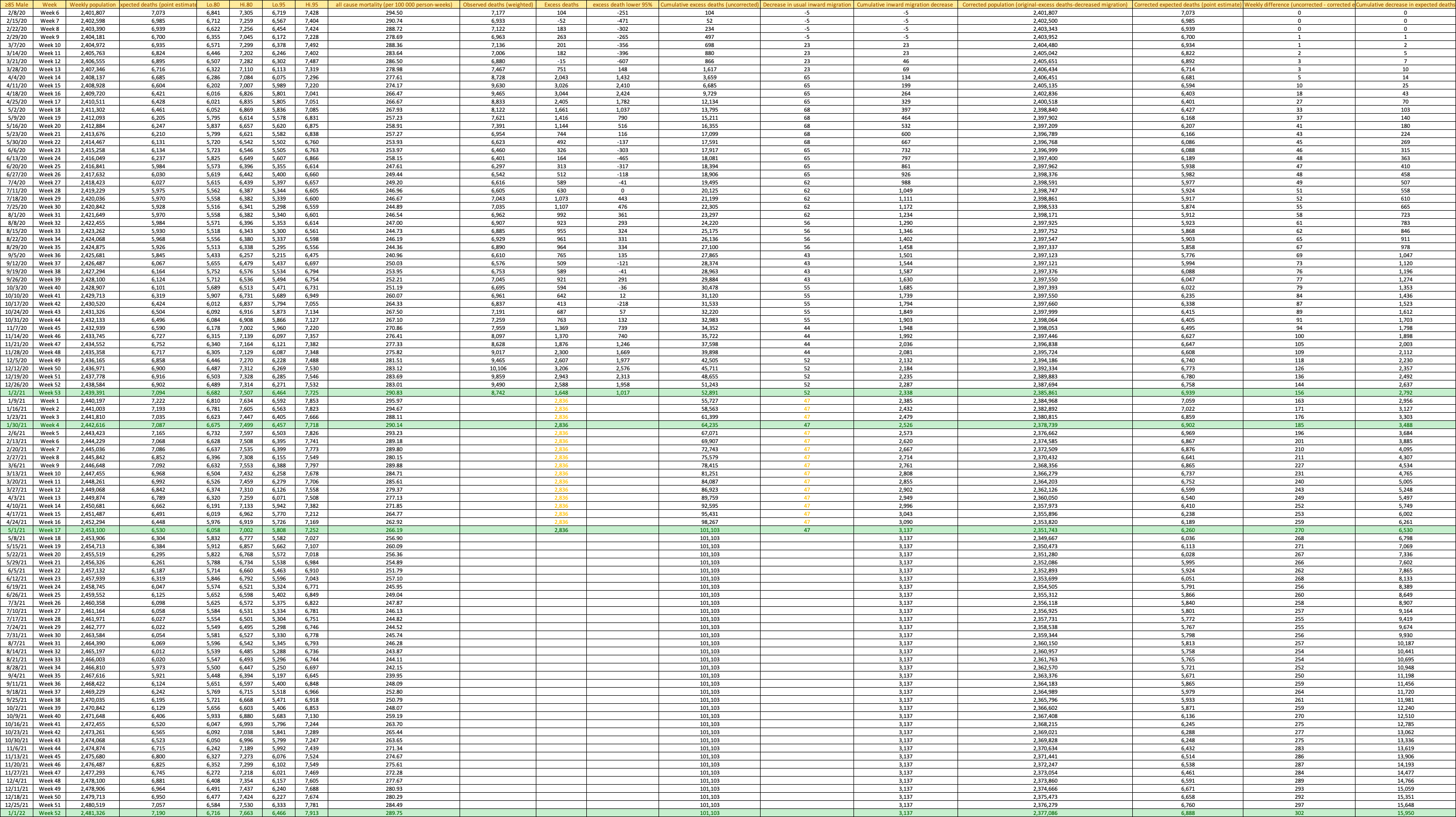

**Table S16.** Weekly population, expected all-cause mortality, excess deaths, population decreases, corrected expected and excess deaths, United States total, Week 6, 2020 through Week 52, 2021.
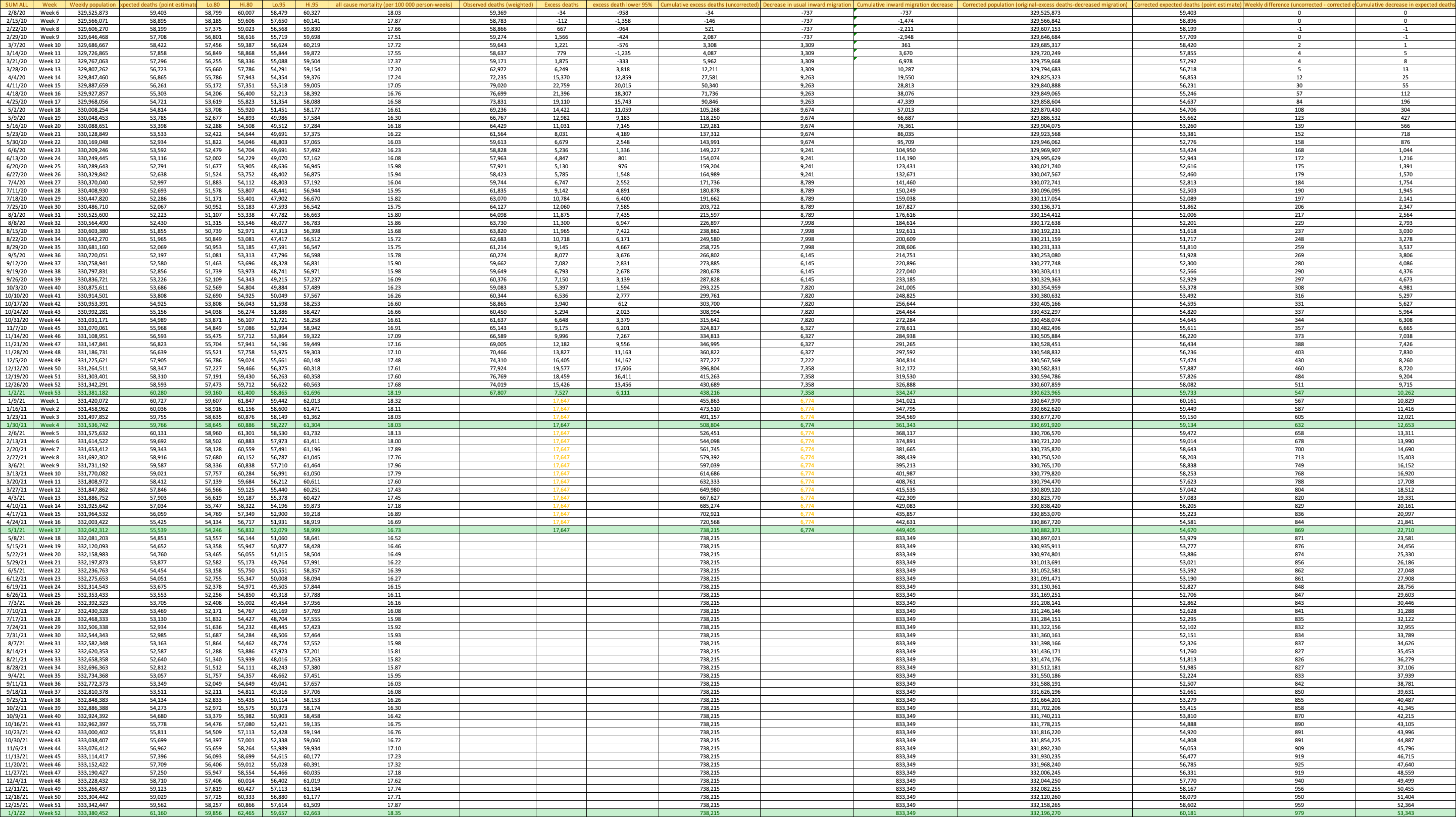
